# Hill numbers at the edge of a pandemic: rapid SARS-COV2 surveillance using clinical, pooled, or wastewater sequence as a sensor for population change

**DOI:** 10.1101/2022.06.23.22276807

**Authors:** Apurva Narechania, Dean Bobo, Kevin Deitz, Rob Desalle, Paul Planet, Barun Mathema

**Affiliations:** Institute for Comparative Genomics, American Museum of Natural History, New York, NY, USA; Section for Hologenomics, The Globe Institute, University of Copenhagen, Copenhagen, Denmark; Department of Ecology, Evolution, and Environmental Biology, Columbia University, New York, NY, USA; Division of Pediatric Infectious Diseases, Children’s Hospital of Philadelphia, Philadelphia, PA, USA; Department of Pediatrics, Perelman College of Medicine, University of Pennsylvania, Philadelphia, PA, USA; Department of Epidemiology, Mailman School of Public Health, Columbia University, New York, NY, USA

**Keywords:** Hill numbers, Information Theory, SARS-COV2, Pangenomes, Population Sweeps, Genomic Surveillance, minhash

## Abstract

The COVID-19 pandemic has highlighted the critical role of genomic surveillance for guiding policy and control strategies. Timeliness is key, but rapid deployment of existing surveillance is difficult because most approaches are based on sequence alignment and phylogeny. Millions of SARS-CoV-2 genomes have been assembled, the largest collection of sequence data in history. Phylogenetic methods are ill equipped to handle this sheer scale. We introduce a pan-genomic measure that examines the information diversity of a k-mer library drawn from a country’s complete set of clinical, pooled, or wastewater sequence. Quantifying diversity is central to ecology. Studies that measure the diversity of various environments increasingly use the concept of Hill numbers, or the effective number of species in a sample, to provide a simple metric for comparing species diversity across environments. The more diverse the sample, the higher the Hill number. We adopt this ecological approach and consider each k-mer an individual and each genome a transect in the pan-genome of the species. Applying Hill numbers in this way allows us to summarize the temporal trajectory of pandemic variants by collapsing each day’s assemblies into genomic equivalents. For pooled or wastewater sequence, we instead compare sets of days represented by survey sequence divorced from individual infections. We do both calculations quickly, without alignment or trees, using modern genome sketching techniques to accommodate millions of genomes or terabases of raw sequence in one condensed view of pandemic dynamics. Using data from the UK, USA, and South Africa, we trace the ascendance of new variants of concern as they emerge in local populations months before these variants are named and added to phylogenetic databases. Using data from San Diego wastewater, we monitor these same population changes from raw, unassembled sequence. This history of emerging variants senses all available data as it is sequenced, intimating variant sweeps to dominance or declines to extinction at the leading edge of the COVID19 pandemic. The surveillance technique we introduce in a SARS-CoV-2 context here can operate on genomic data generated over any pandemic time course and is organism agnostic.

**One-Sentence Summary:** We implement pathogen surveillance from sequence streams in real-time, requiring neither references or phylogenetics.

**Main Text:** The COVID-19 pandemic has been fueled by the repeated emergence of SARS-CoV-2 variants, a few of which have propelled worldwide, asynchronous waves of infection(1). First arising in late 2019 in Wuhan, China, the spread of the D614G mutation led to sequential waves of Variants of Concern (VOC) about nine months later, significantly broadening the pandemic’s reach and challenging concerted efforts at its control (2). Beta and Gamma variants drove regional resurgences, but Alpha, Delta and Omicron occurred globally (3)(4). The advent of each variant led to the near extinction of the population within which it arose (5). The architecture of this pandemic is therefore marked by periods of transition, tipping a population towards an emerging variant of concern followed by its near complete sweep to dominance.

At the pandemic’s outset, epidemiological work was focused on transmission networks, but SARS-CoV-2’s high rates of infection quickly outstripped our ability to trace it(2). When it became clear that even focused global efforts would only characterize a fraction of infections, researchers turned to phylodynamic approaches to understand SARS-CoV-2’s population structure(6)(7). Genomics was at the center of this effort. Rapid sequencing and whole genome phylogeny updated in quasi real time enabled epidemic surveillance that was a few weeks to a month behind the edge of the pandemic curve(8). In a crisis of COVID-19’s scale and speed, eliminating this analysis lag can mean the difference between timely, reasonable public health response and failure to understand and anticipate the disease’s next turn.

Phylodynamics is predicated on genetic variation. Without variation, phylogenetic approaches yield star trees with no evolutionary structure. The high mutation rate among pathogens, especially among RNA viruses like SARS-CoV2, ensures the accumulation of sufficient diversity to reconstruct pathogen evolutionary history even over the relatively short time scales that comprise an outbreak. But as a genomic surveillance technique, phylodynamics is costly. Tools like Nextstrain align genomes, reconstruct phylogenies, and date internal nodes using Bayesian and likelihood approaches(9). These techniques are among the most computationally expensive algorithms in bioinformatics. Intractable beyond a few thousand sequences, phylodynamic approaches must operate on population subsamples, and subsamples are subject to the vagaries of data curation. More importantly, phylodynamic approaches are yoked to references. Most techniques are ill-equipped to respond to evolutionary novelty. We argue that genomic surveillance should herald the appearance of previously unseen variants without having to resort to comparison with assembled and curated genomes, and the lag between variant discovery and a database update is often months. Surveillance is currently hamstrung by the historical bias inherent to marker-based analysis. The existing pandemic toolbox therefore lacks unbiased approaches to quickly model the population genomics of all sequences available.

We propose a method that summarizes the temporal trajectory of pandemic variants by collapsing each day’s assemblies into a single metric. In the case of pooled or wastewater sequence, this same metric is repurposed to measure survey sequence compression across days. Our method does not subsample, perform alignments, or build trees, but still describes the major arcs of the COVID19 pandemic. Our inspiration comes from long standing definitions of diversity used in ecology. We employ Hill numbers (10)(11), extensions of Shannon’s theory of information entropy(12). Rather than using these numbers to compute traditional ecological quantities like the diversity of species in an area, we use them to compute the diversity of genomic information. For example, we envision each unique k-mer a species and each genome a transect sampled from the pan-genome. Applying Hill numbers in this way allows us to measure a collection of genomes in terms of genomic equivalents, or a set of sequence pools as the effective number of sets. We show that tracing a pandemic curve with these new metrics enables the use of sequence as a real time sensor, tracking both the emergence of variants over time and the extent of their spread.

## Implementation

To measure the information diversity of a population, we use ecological definitions that condense the influence of millions of genomes or terabases of raw sequence into a single value. In ecology, the effective number of species is a quantity used to provide an intuitive and simple metric for comparing species diversity across environments. Hill(10) defined the effective number of species of order q as

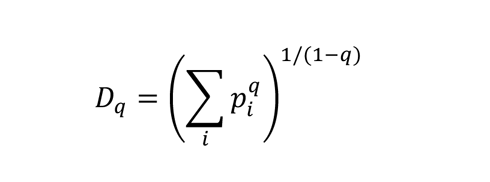

where *pi* is the frequency of a particular species *i*. As q tends towards one, the limit of this equation invokes Shannon’s concept of generalized information entropy. The exponent of the Shannon entropy yields the effective number of species, a quantity commonly referred to as the Hill number.

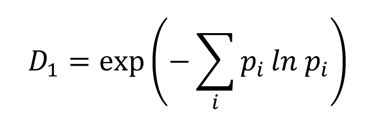

Hill numbers are best conceptualized as the number of equally abundant species required to recapitulate the measured diversity in a sample. The more diverse a sample, the higher the Hill number. The Hill number is not unique to ecological analysis. For example, in Natural Language Processing (NLP), this same quantity is termed perplexity, a key measure in the evaluation of language models(13).

While species Hill numbers are helpful for understanding the nature of any given sample, ecologists are often interested in comparisons across samples and communities. The motivation is to assess experimental design and/or to gauge the piecemeal complexity of an ecosystem. The beta diversity, a measure of differentiation between all local sites, extends species Hill numbers to Hill numbers of communities. Ecologists use the effective number of communities to understand the competing effects of their sampling and the innate diversity of their transects(14)(15)(16). The Hill number based on beta diversity is given as

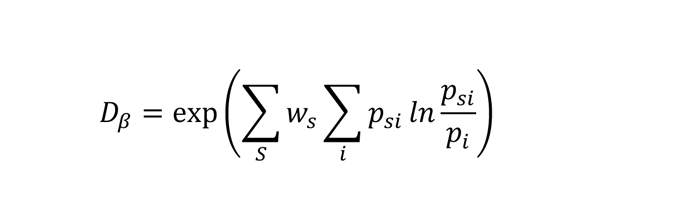

an expression that incorporates the Kullback-Leibler divergence(17) to yield the effective number of communities. Here, *psi* is the frequency of a particular species *i* in a particular sample *s, pi* is the frequency of a particular species *i* across all samples, and *ws* weighs all observations in sample *s* relative to all individuals censused in the experiment.

The effective number of communities measures the compressibility of the samples. If the samples share many species and if individuals are evenly distributed across these species, the samples essentially collapse, yielding a lower number of effective communities. If the samples are highly diverse and the individuals scattered among these divergent organisms, the effective number of communities approaches the number of communities sampled (Figure 1B). This interpretation of the beta diversity relies heavily on information theoretic principles, reflecting the broad and powerful implications of Shannon’s original theory of entropy and communication(12).

**Figure 1.**
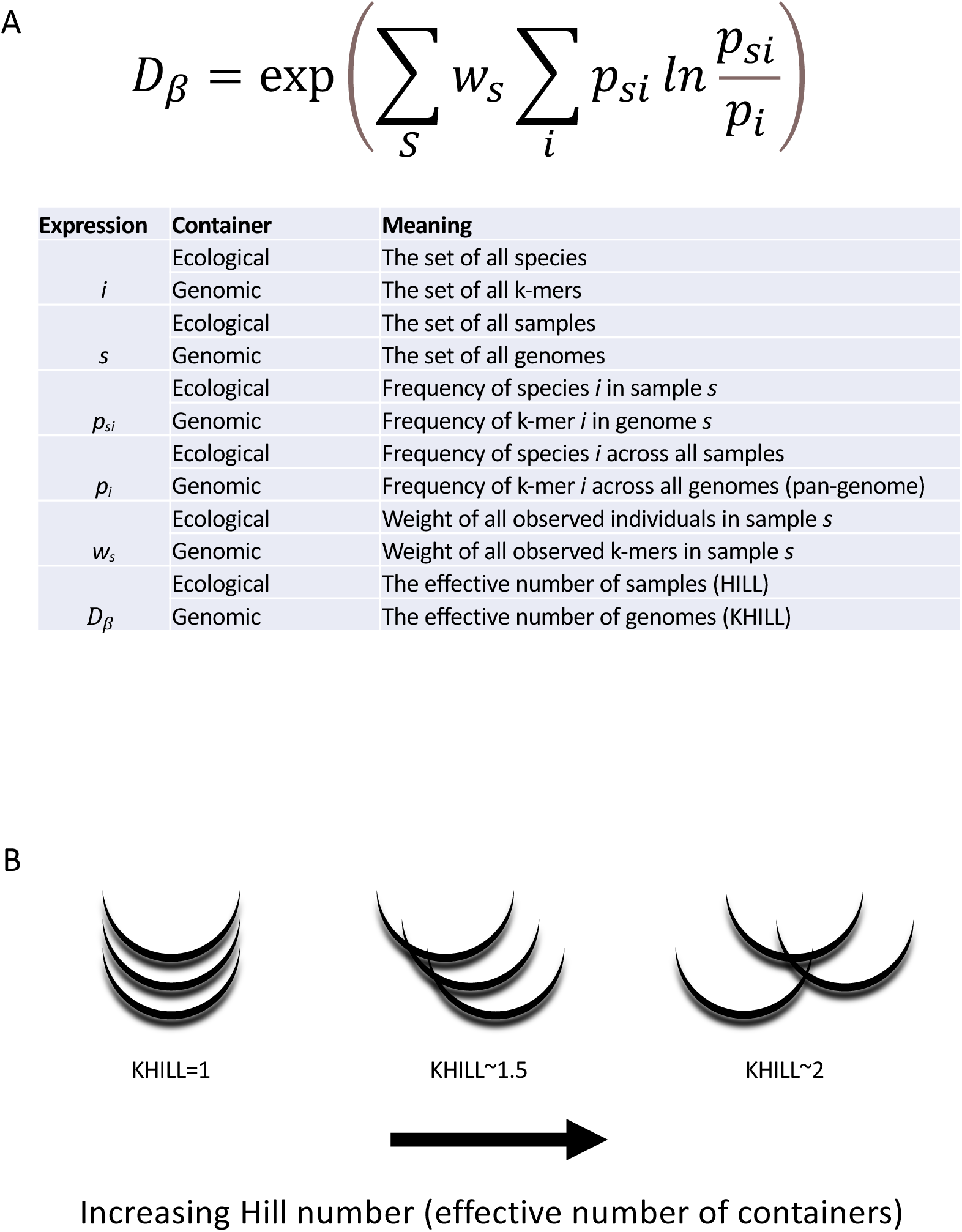
The KHILL metric. We adapt ecological definitions for beta diversity to calculate the effective number of genomes (KHILL) in any set. We explicitly describe the analogy in terms of variables in the KHILL expression (Panel A). Genomes with overlapping k-mer identities and similar k-mer frequencies will tend to have a lower KHILL (Panel B).

In the ecological framework we have described, the sample or community functions as a container for observations of species and their frequencies. However, the container can be anything.

Here, we reframe these equations to describe a system where the containers are either discrete, clinical genomes or pools of samples. Pools can be combined at the clinic or gathered downstream in wastewater. The species within these containers are enumerated strings of information or k-mers. For clinical sample genomes, the effective number of communities becomes the effective number of genomes, or genome equivalents. For sequences pooled by day, this same quantity becomes the effective number of days. Regardless of container, we term this new quantity KHILL. If we consider a set of identical sequence, KHILL will reduce to 1, whereas in a set of sequence with no information overlap, KHILL will achieve its maximum, the number of sets considered. The analogy is made explicit in Figure 1.

Equipped with this new metric of genome diversity, we can track population dynamics across any axis of change. For clinical genomes over a defined time course, a stable community of species subtypes may experience a disturbance that signifies the emergence of a new variant. If selection favors this variant, the population will pass through a KHILL peak where genome equivalents find a local maximum, and stable subtypes will coexist in almost equal measure with the newcomer. This local maximum should erode if the new variant sweeps through the population. When prior subtypes are driven to extinction and a single variant pervades, we expect a local minimum for KHILL or the effective number of genomes (Figure 2A). For sequence pools, we instead compare the most recent day’s survey sequence to sequence from all prior days. In this case, our containers are not discrete clinical genomes. Instead we calculate KHILL from infection pools. These pairwise comparisons of the most recent day’s k-mer pool to all prior days is similar in spirit to autocorrelation, but our metric is KHILL, the information diversity compressed into the effective number of days. If recent pooled sequence diverges from past sequence, we expect a bend in the KHILL curve describing a change in sequence compression. For pairwise comparisons, KHILL will vary between 1 and 2 (Figure 2B). We mark significant changes in the time course using Bayesian Change Point detection(18).

**Figure 2.**
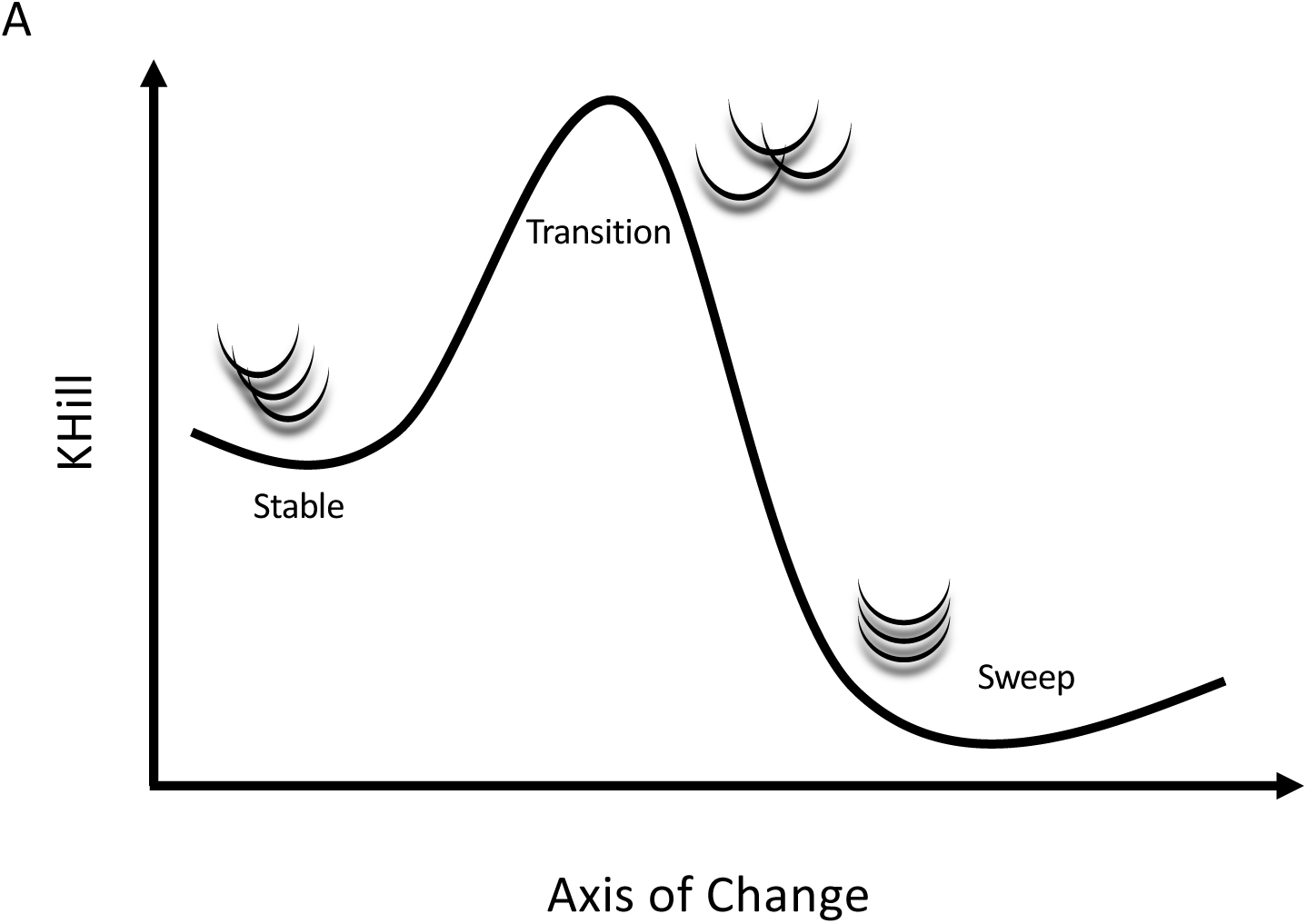
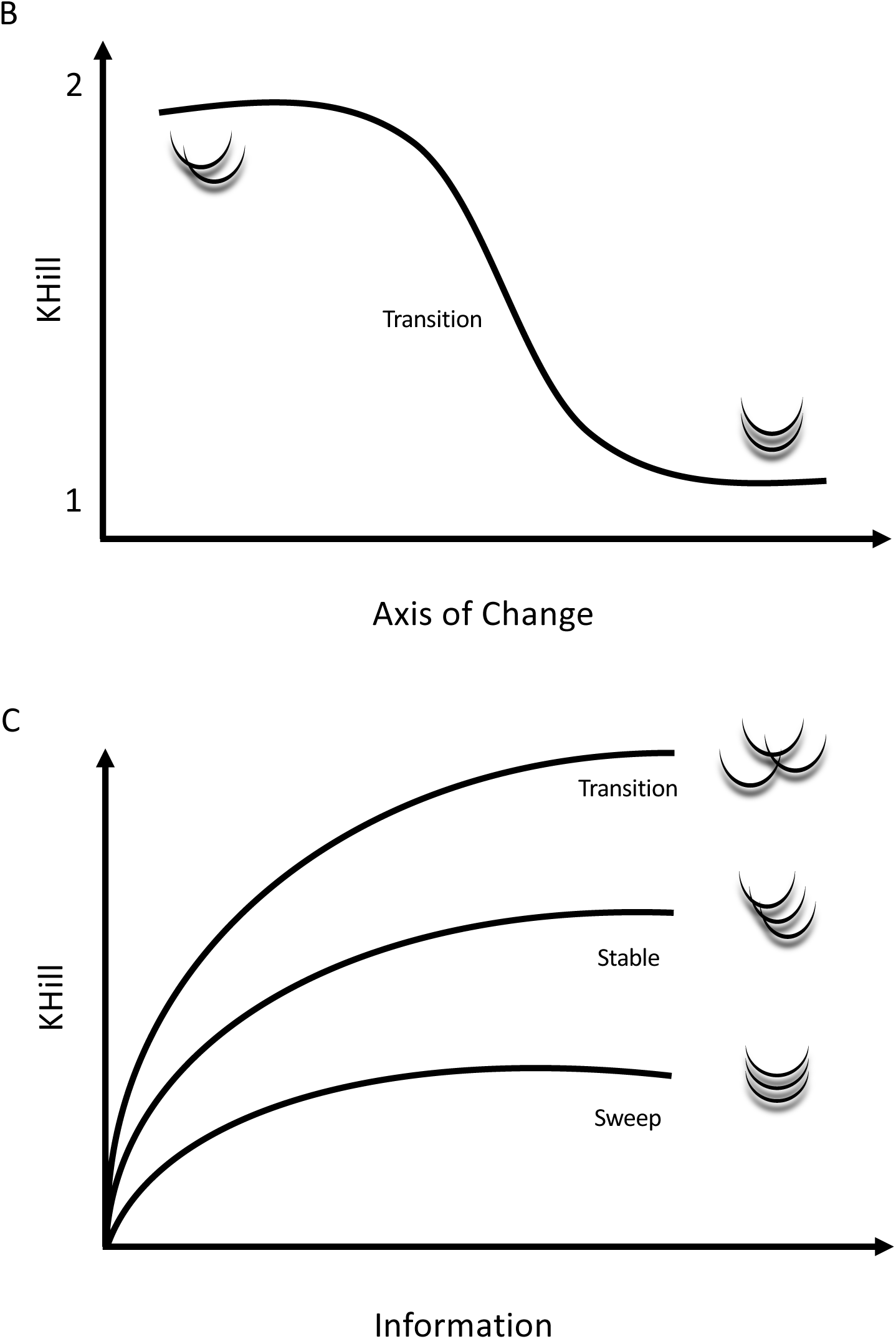
KHILL cartoons. In a population genomic context, KHILL can be used to track changing clinical genomic complexity over time and/or space. We expect that in a pandemic, a Variant of Concern will increase the information diversity of a stable population. If this transition is favored, the variant will sweep the population (Panel A). In a pooled or wastewater context, KHILL can function as a measure of compression of unassembled, raw sequence across days, with a drastic change revealing the potential arrival of a new variant (Panel B). Genomes sampled from a transitioning population will exhibit higher KHILL than a population that has settled into a dominant variant (Panel C).

A further application of this approach is to pangenomes, the accrued genomic content of any given species(19). Because the KHILL metric functions as a measure of container equivalents independent of functional content, it can bring new clarity to how pangenomes change in space and time. The state of the art in pangenomes is wedded to genes and/or alignments. Phylogenomic methods calculate orthologs(20), and pangenome graphs fork alignments along multiple, disparate paths(21). While a number of these methods have achieved significant speed (22), KHILL obviates the need for phylogenetics entirely by calculating information theoretic quantities on containers of k-mers. We think not in terms of genes, but in terms of unique strings of information. As we accumulate information, the KHILL curve of a transitioning population will exhibit information diversity of a higher steady state than the simpler population left in the wake of a selective sweep (Figure 2C).

Since the KHILL approach operates on containers of unaligned strings our main computational challenge is to reduce the number of string comparisons without sacrificing sensitivity. We find that a string sketch(23), as implemented in programs like MASH(24), adequately reflects calculations made on the whole population. We need only a fraction of available k-mers to carry each container’s signal. Using a bottom-k sketch (25) of hashed strings, we accelerate KHILL calculations by many orders of magnitude.

Our approach is unusual in that we are not looking for evidence of specific genetic events. With sketched strings of unaligned data, we sacrifice most of the products of modern bioinformatics: the discovery of mapped genome variation like alleles in particular genes, indels in non-coding regions, or genome-scale structural rearrangement. But we gain a fast, intuitive metric that summarizes the contributions of both micro-variation on the mutational scale, and macro-variation such as the presence/absence of genomic elements.

## Results and Discussion

The United Kingdom is a model for COVID-19 genomic surveillance(26). The COVID-19 Genomics UK (COG-UK) consortium has accumulated more genomes and more metadata than any other regional health organization. Figure 3 shows how the seven-day moving average of the 22.2 million UK cases reported as of July 15, 2022 (panel A) has spikes coincident with the emergence of the UK’s three main variants of concern: Alpha, Delta and Omicron (panel B). Panels A and B rehash the accepted epidemiological approach, case counts augmented by phylogenetic annotation. Using the complete set of 2.5 million UK genomes sequenced from these reported cases, we add panel C, a single KHILL value calculated for each day. With the emergence of each variant, KHILL rises until it reaches a local maximum where sequenced genomes are evenly distributed between the old variant and the new.

**Figure 3.**
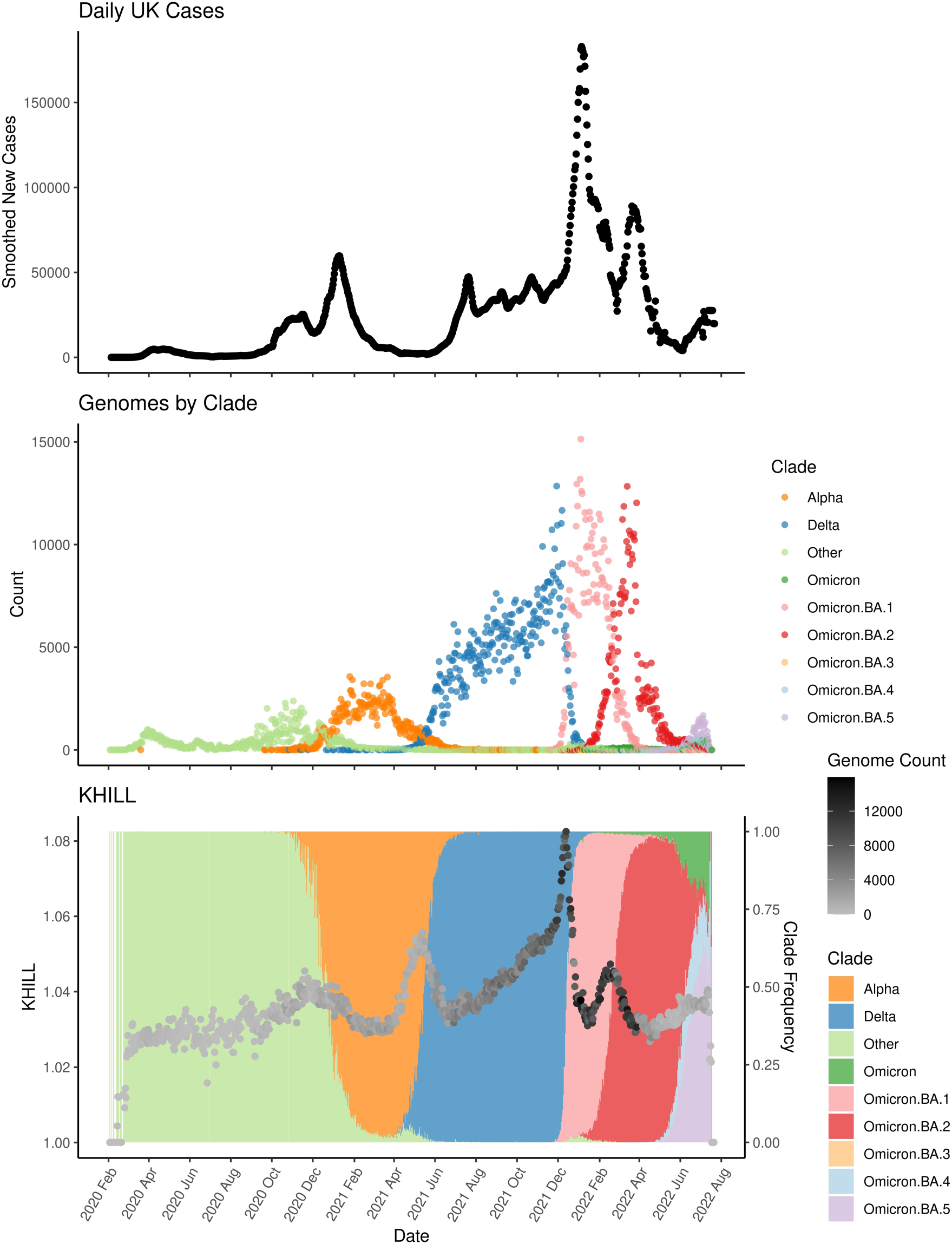
The United Kingdom (UK) COVID-19 pandemic. We show new case burden (Panel A) and gross phylogenetic classification (Panel B) for the UK pandemic since its inception. The three distinct KHILL peaks signal the arrival of Alpha (November 2020), Delta (May 2021), and Omicron (November 2022) well before each variant’s spike in cases (Panel C).

Against a background of The UK’s dominant variant waves, these peaks mark clear transitions in the pandemic. Notably, the stark peaks that presage the emergence of Delta and Omicron occur well before each variant’s burden of cases. In May 2021, public measures like masking and travel restrictions suppressed new infections, but it is in this month that we observe the KHILL peak signaling the arrival of Delta. The changing complexity of the viral population is reflected in this curve of daily genome equivalents regardless of the volume of cases and regardless of the variant. Because the expression for the effective number of genomes is weighted, days with just tens of sequences scale with days that may have tens of thousands.

In the UK, each population transition signified by the three KHILL peaks, were harbingers of a selective sweep. A peak’s ascent reflects the accumulating momentum of a variant of concern, while the descent suggests this new variant’s primacy. Alpha swept the population in March 2021 (27), Delta by July 2021 (28), and Omicron by January 2022 (29). In each case, KHILL settles into a local minimum of about 1.03 effective genomes highlighting the near clonality of the viral population after a variant achieves dominance. Rising KHILL values within the Delta and Omicron waves suggest that we also detect accruing diversity within the Delta (mixing of AY.4, B.1.617.2, and AY.4.2) and Omicron (BA.1 and BA.2) lineages over time. Though evidence of evolving subvariants requires a deeper sketch (Supplemental Figure 1), this within wave signal tracks the information complexity of steady evolution following a selective sweep.

We performed the same type of analysis for both the United States and South African pandemics. Though we see regional differences in terms of which variants were dominant at which times, in every case, the KHILL curve tracks the arrival of prevailing variants. Unlike the UK, the start of the US pandemic is variably complex with near simultaneous transmission events from around the world (30)(31) manifesting as a noisiness in KHILL (Supplemental Figure 2). The introduction of Alpha flattens this diversity. In South Africa, the Beta variant achieved a higher share of infection than in either the UK or the US (32). Though sparse genomic sampling results in a stochastic KHILL curve, in SA we observe clear peaks demarcating the transitions between Beta/Delta and Delta/Omicron (Supplemental Figure 3).

As SARS-CoV-2 surveillance transitioned from patient genomes gathered in the clinic to pooled sequence accumulated in sewage, we extended the KHILL statistic to operate on sequence dissociated with individual infections. Sequence of this type is necessarily unassembled. We show that since KHILL relies only on sketched strings, reads alone produce a curve that mirrors the pattern we see for all assemblies. For the curve in Supplemental Figure 4, we simulated 250,000 reads from each genome of a random sample of 100 genomes selected daily over the course of the UK pandemic.

Pooled sequence requires that we dissolve the boundaries between these discrete infections. Rather than sampling individual infections, we pool a population. To model these pools, we joined and shuffled simulated reads generated for each day’s infections, retaining only 400,000 as a kind of daily signature or sensor read for the overall state of the region. The conceptual shift here is in the nature of the container. From containers of discrete genomes we transition to days of pooled infections. Figure 4 features three curves chosen to highlight KHILL dynamics for the three global variants. In each case, we query the curve’s endpoint (most recent time point) against sequence surveyed from all days prior. The emergence of each variant is marked by a clear, sometimes precipitous, downturn in the value of KHILL, creating at least two starkly different populations: days before the variant appeared and days after. In the case of Delta and Omicron, shadows of the previous variants are also clearly visible. For statistical rigor, we apply Bayesian Change Point analysis to tag significant transitions and note that these detected points correspond to known population dynamics. Notably, posterior probability peaks within the Omicron transition correspond to Omicron subvariants (Figure 4B).

**Figure 4.**
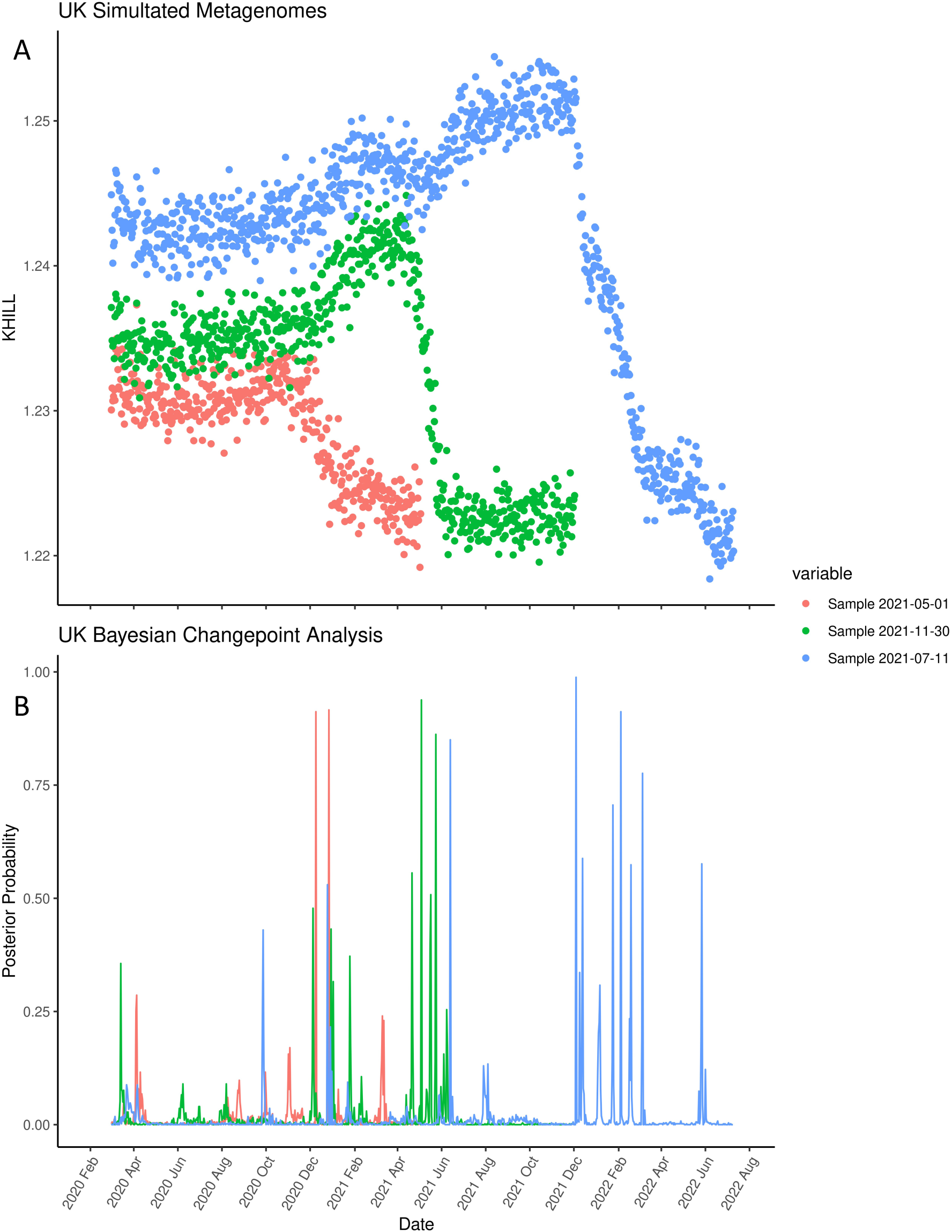
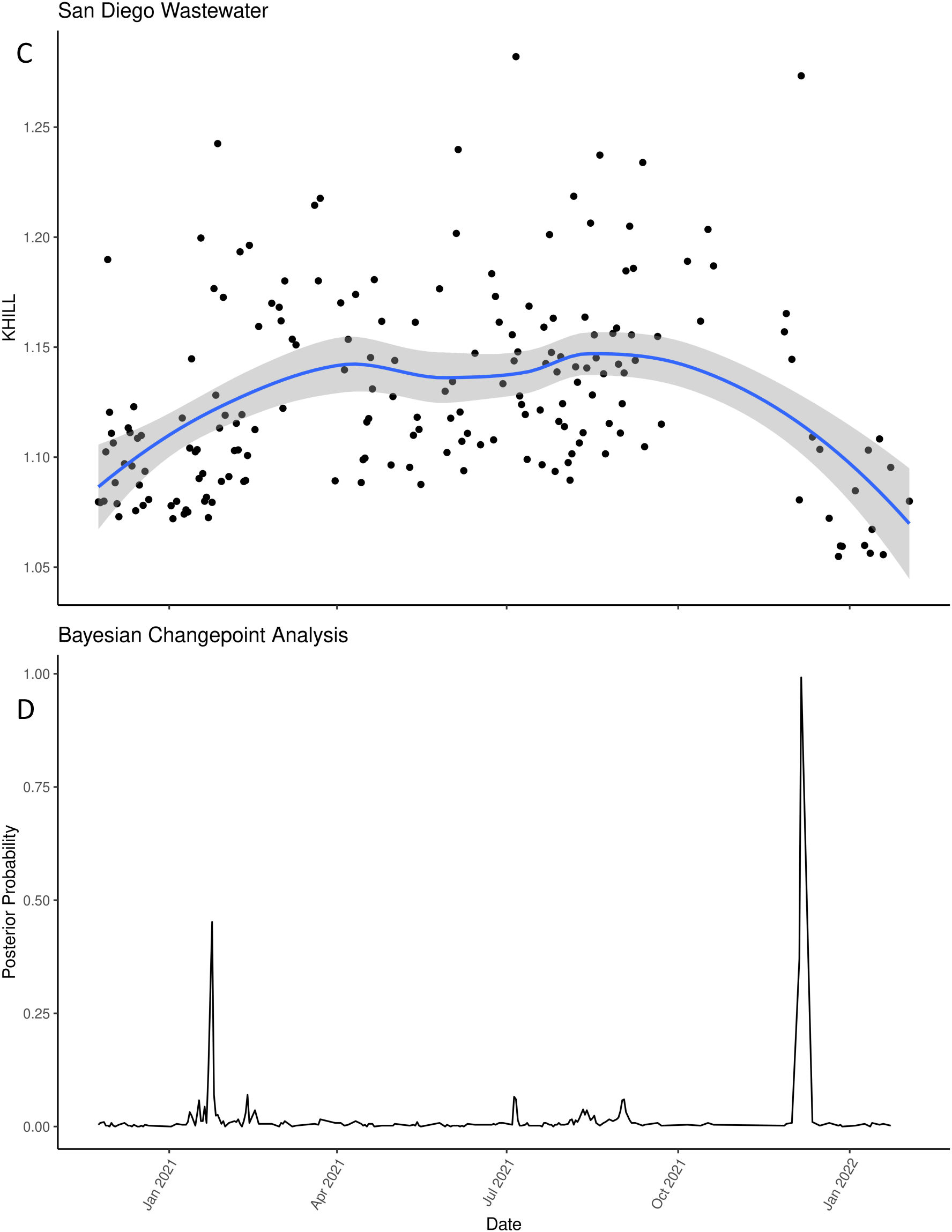
SARS-CoV-2 in raw, pooled sequence. We extract three curves from an autocorrelation analysis of simulated reads from the UK pandemic. The curves chosen focus on the three major variant transitions: alpha, delta and omicron. In each case, the large downward swing marks turnover from one type to another (Panel A). We show that these transitions are significant using Bayesian Changepoint Detection (Panel B). San Diego wastewater data shows similar – albeit noisier – transitions between variants. Comparison of raw sequence from a day dominated by omicron to all prior days shows clear (Panel C) and significant (Panel D) separation between alpha/epsilon and delta/omicron.

Clearly, simulated sequence pools are amenable to KHILL surveillance analysis. But will the technique work for real data? In principle, wastewater is an infection pool. Changes in wastewater information diversity should expose new variants. In Figures 4C and D we show that despite the noise inherent to wastewater data, we discern two clear transitions in the San Diego pandemic (33): the rise of alpha in January of 2021, and the rise of omicron that December. Because KHILL measures change in terms of k-mer representation and frequency across days, it is sensitive to the technical errors and noise characteristic of metagenomic sequencing in a way that marker-based analysis is not. To screen the noisiest time points, we calculated the alpha diversity of k-mers sampled from each day and kept only those days within one standard deviation of the mean (Supplemental Figure 5). This alpha diversity screen tightened our curve significantly, but the emergence of delta, which is known to have dominated San Diego infections by July 2021, remains hidden. Karthikeyan et al (33) use references to squash noise by forcing alignment, but alignment limits novelty detection through comparison to the known. Given comparable sequence coverage and depth across days, KHILL can eliminate the need for references, quicken the pace of discovery, and capture incipient novelty likely to be lost to marker-based analysis. Computing KHILL from pooled or wastewater reads realizes our ambition of making streamed sequence a kind of online sensor (34) that can aggregate signal directly and in real time and the very edge of a pandemic.

To illustrate how traditional methods lag in their detection of variant changes in a population, we revisit the UK KHILL curve, focusing on the emergence of BA2. Figure 5A shows the curve overlayed on version 1.2.101 Pangolin annotations. This version was released on January 28^th^, 2022. Prior to this release, it is already clear that a steady increase in the effective number of genomes is underway, culminating in a peak just a few weeks later. But the 1.2.101 Pangolin annotation conflates this additional heterogeneity into a single annotation for B.1.1.529. About one month later, in panel B we see that the KHILL trace anticipated the arrival of a new Omicron subvariant, BA2. BA2 entered the lexicon too late to offer useful classification for what was clearly an emerging variant as early as January 1^st^, 2022. The BA2 classification seems to trail the BA2 KHILL peak by a full month and the beginning of the BA2 curve by two months.

**Figure 5.**
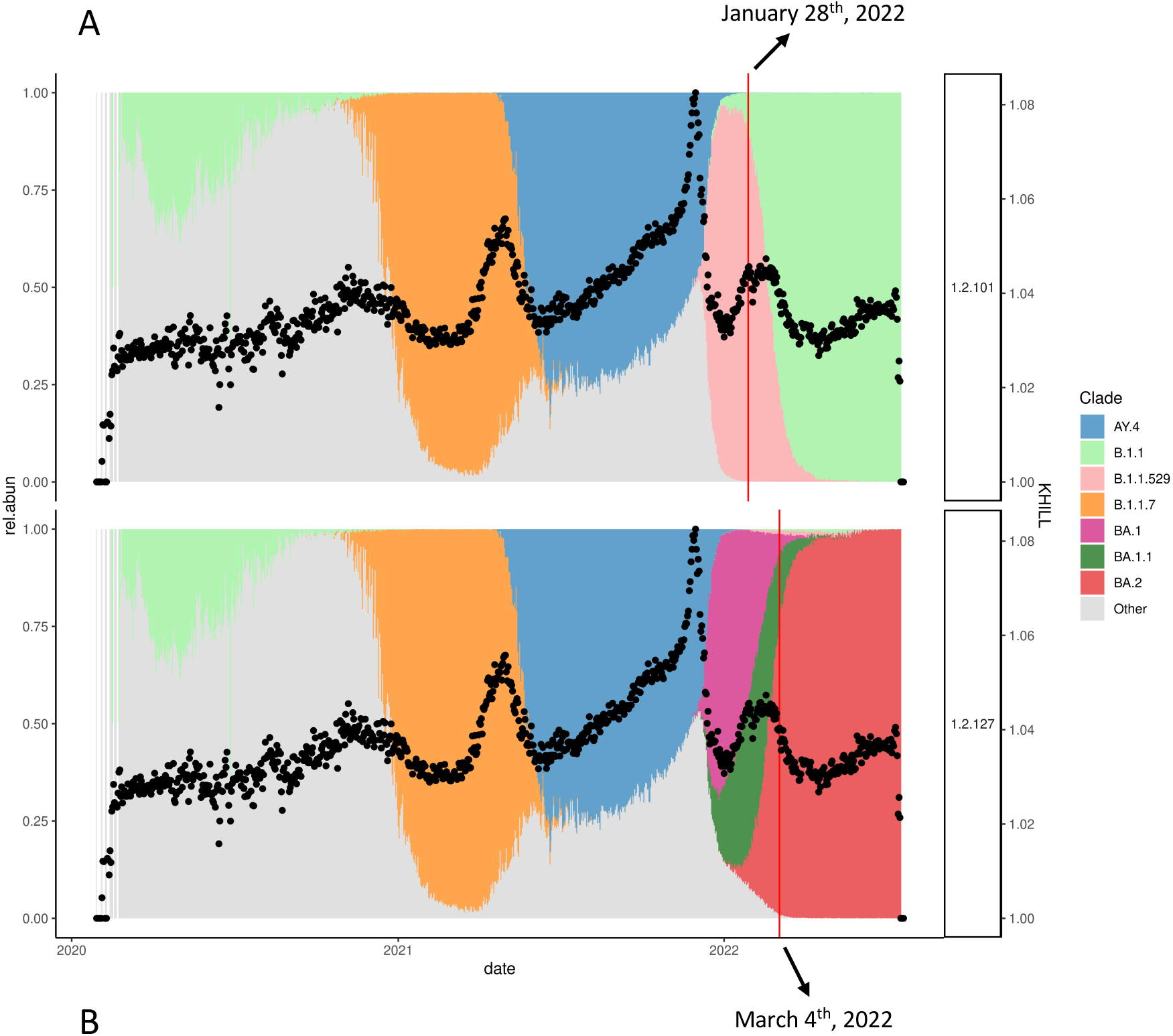
KHILL anticipates new variants. We show that in New York State clinical isolates, a KHILL peak around December 2022 forecast the emergence of the new Omicron subvariant, XBB. But this variant was not properly classified until two months later, well after XBB had saturated the population (Panels A and B). We show that in the UK population, BA2’s classification was similarly delayed and only properly fixed into the Pangolin databases well after KHILL indicated BA2’s population suffusion (Panels C and D).

KHILL was picking up something new well before BA2 was fixed into the databases offering an immediacy that phylogenetic, reference-bound, and annotated techniques like Pangolin lack. KHILL is sensitive and capable of monitoring shifts in the viral population despite the handful of changes that distinguish each variant. To better understand the number and type of phenomena we can detect with KHILL, we simulated blocks of change across 100 30kb genomes varying each population from one base to 500 contiguous bases across several classes of mutation including insertion, deletion, duplication, and transposition. (Supplemental Figure 6)(35). For SARS-CoV-2, blocks of 1 to 5 bases are biologically relevant. Within this scope, we see that 10 changes raises the effective number of genomes to about 1.05, a value that mirrors biological reality. As we would expect, any change that alters the population of k-mers (e.g., insertion) results in greater KHILL values than change that simply alters the existing k-mer counts (duplication, transposition). With frequent insertion or change over long block sizes, we see a KHILL that approaches the number of genomes simulated, a range that conveys the power of the KHILL metric to track both subtle and gross genomic change.

Our approach is fast, straightforward and ideally suited to durable but nimble real-time genome surveillance. We calculate Figure 2C on all UK genomes available in 1800 CPU hours on low memory cores. If we decrease the sketch rate by an order of magnitude, we can calculate the same curve in just 180 CPU hours without losing signal from any of the UK’s major variant transitions (Supplemental Figure 1). The omicron autocorrelation curve in Figure 3A is similarly fast. We calculate all 860 required KHILL values in 2 CPU hours. We can therefore easily append daily KHILL points to the growing epidemiological time series. Recent studies struggle to process all available genomes in heavily sampled regions(36). Curation can be a viable strategy. In Supplemental Figure 7, we show that genome wide nucleotide diversity (π) of 100 randomly selected daily sequences over the UK pandemic mirrors the KHILL curve but distorts its amplitude. But the time required to calculate alignments for π makes this an impractical solution at the scales we’ve proposed here.

Though we prescribe no thresholds for marking the emergence of a new variant, our technique affords public health institutions the opportunity to create actionable policy based on a simple, quantitative measure. Moreover, we show that the first derivative of the KHILL curve tracks new variants as deviations from a critical point, providing another potential metric to trigger policy changes. During a KHILL upswing, the increasingly complex population is marked by an accelerating slope, which then flattens and decelerates until the downswing settles into a variant sweep (Supplemental Figure 8).

Finally, KHILL not only distinguishes the various states of genome populations along an axis of change, but also quantifies exactly how much genome diversity emerges during a transition, and how much is lost in a sweep. In other words, KHILL not only responds to a changing population, but also integrates the effects of genetic distance, merging two primary properties of pan-genomes. This is true of both the clinical and pooled samples.

Applied to genomes that comprise species level complexes, we believe that KHILL accumulation curves can function as a new kind of comparative genomics realized without the usual computational burden. For example, in Figure 6 we show that, in the UK population, as we accumulate Delta and omicron genomes, we plateau at about 1.05 genome equivalents. Alpha and Omicron BA1 asymptote at about 1.03. Higher Delta genome diversity agrees with conclusions drawn from slower phylogenetic methods(37). We quickly capture this species level trait using the differential saturation rate of a metric that, for any set of genome sequences, collapses all genomic diversity into a single point.

**Figure 6.**
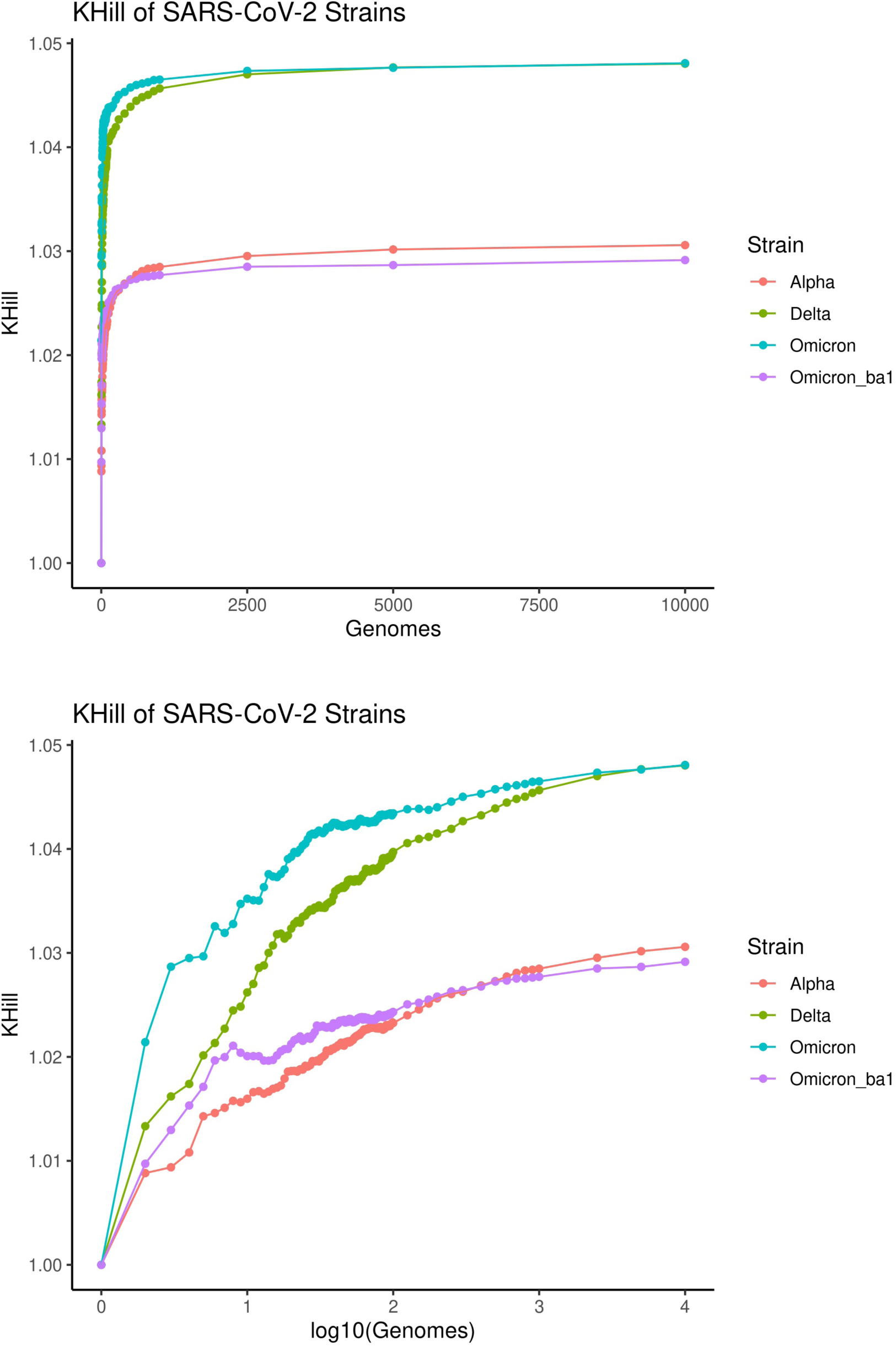
SARS-CoV-2 comparative genomics. We randomly sampled 10,000 Alpha, Delta and Omicron genomes from the UK pandemic. As genomes are randomly accumulated, information diversity accrues. We show that Delta and Omicron converge on similar information diversity, but that Delta is more complex. Both are far more diverse than either Alpha or Omicron BA1 alone, an observation confirmed by phylogenetic methods on far fewer genomes.

KHILL is ideal for population genomics, but the approach is also relevant for biological problems at other evolutionary scales. The comparative approach we highlight here might help illuminate the species concept itself. With genomic containers, how many genome equivalents should a legitimate species contain? Can a comparative genomics of KHILL accumulation curves complement or even enhance traditional comparative genomic methods based in phylogenies or networks? Moreover, containers of pooled sequence or shotgun metagenomes could be used to understand the changing diversity of environmental samples in both time and space. We show the potential in this type of analysis in the autocorrelations selected for Figure 3.

In this work, we use Hill numbers and the notion of container equivalents for genomic surveillance. The response to the COVID-19 pandemic has resulted in some of the richest biological datasets in history. Our technique is positioned to stream all this data. The state of the art is burdened by alignment, a reliance on known references, and the lag characteristic of retrospective databases. We do not stream through markers. Our goal is to signal population change regardless of taxonomy. We see great value in a simple measure that conflates the evolutionary nuance of phylodynamics without sacrificing a pandemic’s actionable signals of transition and sweep. The method we describe here can accommodate the massive genomic datasets of future outbreaks regardless of organism, and perhaps signal when phylogenetics are needed. KHILL is a new angle on the COVID19 pandemic, a technique that takes us closer to the edge of one of the greatest health care challenges of our time.

## Data Availability

All data produced are available online at:
https://www.cogconsortium.uk
https://www.gisaid.org

## Acknowledgements

We thank Daniel Hooper for his contributions to this effort.

## Funding

National Institute of Allergy and Infectious Diseases of the National Institutes of Health under award number R01AI151173 (BM)

Centers for Disease Control 75D30121C11102/000HCVL1-2021-55232 (PP)

Women’s Committee of CHOP (PP)

## Author Contributions

Conceptualization: AN

Methodology: AN

Investigation: AN, DB, KD, RD, PP, BM

Visualization: DB, KD

Supervision: AN, PP, BM

Writing (original draft): AN

Writing (review and editing): AN, PP, BM

## Data and materials availability

All data sources are available in the main text or the supplementary materials. Code can be found here: https://github.com/narechan/khill.

## Supplementary Materials

Supplemental methods

Table S1

Figures S1 to S8

## Supplemental Methods

### UK Clinical Genomes

Genomes and metadata were downloaded from the Covid-19 Genomics UK Consortium (COG-UK) https://www.cogconsortium.uk/. A custom R script (R version 4.1.3) was used to read the genomes and metadata into memory and sort genomes into one directory per day. We used collection dates to sort into daily directories. A total of 2,794,151 genomes were analyzed. The KHILL program was run on each directory with the following parameters: -k 19 -m 1 -n 100 -s 1e99 -p 6. To calculate daily cases, we downloaded covid cases data from Our World In Data https://ourworldindata.org/covid-cases. The csv file was loaded into R and filtered to include only data from the UK. To assign each genome to a clade, we used Pangolin for each genome, https://github.com/cov-lineages/pangolin (pangolin version 4.1.2; data version 1.12). The scorpio call output was used, and we summarized the clade assignments with the modifications shown in Table 1 below. Plots were made with R and ggplot2 (version 3.3.5)

### USA Clinical Genomes

Genomes and metadata were downloaded from the Covid 19 Data portal (https://www.covid19dataportal.org/). A table of metadata for each genome were downloaded from the web interface. The search parameter was (country:“USA”) AND (coverage:[“95” TO “100”]). The accessions were fed into the CDP-File-Downloader (https://www.covid19dataportal.org/bulk-downloads), and genomes were downloaded and put into directories (one directory per day). A total of 1,926,292 genomes were analyzed for the USA using the same methods outlined for the UK above.

### SA Clinical Genomes

Genomes were downloaded from GISAID EpiCov https://www.epicov.org/ We searched for Location = “Africa / South Africa”, selecting “Complete” and “Collection Date Complete”. We downloaded genomes directly from the web interface (selecting Download and then input for “Input for the Augur pipeline”). Genomes were sorted into directories (one directory per day). A total of 32,623 genomes were analyzed for South Africa using the methods outlined for the UK above.

**Supplemental Table 1:**
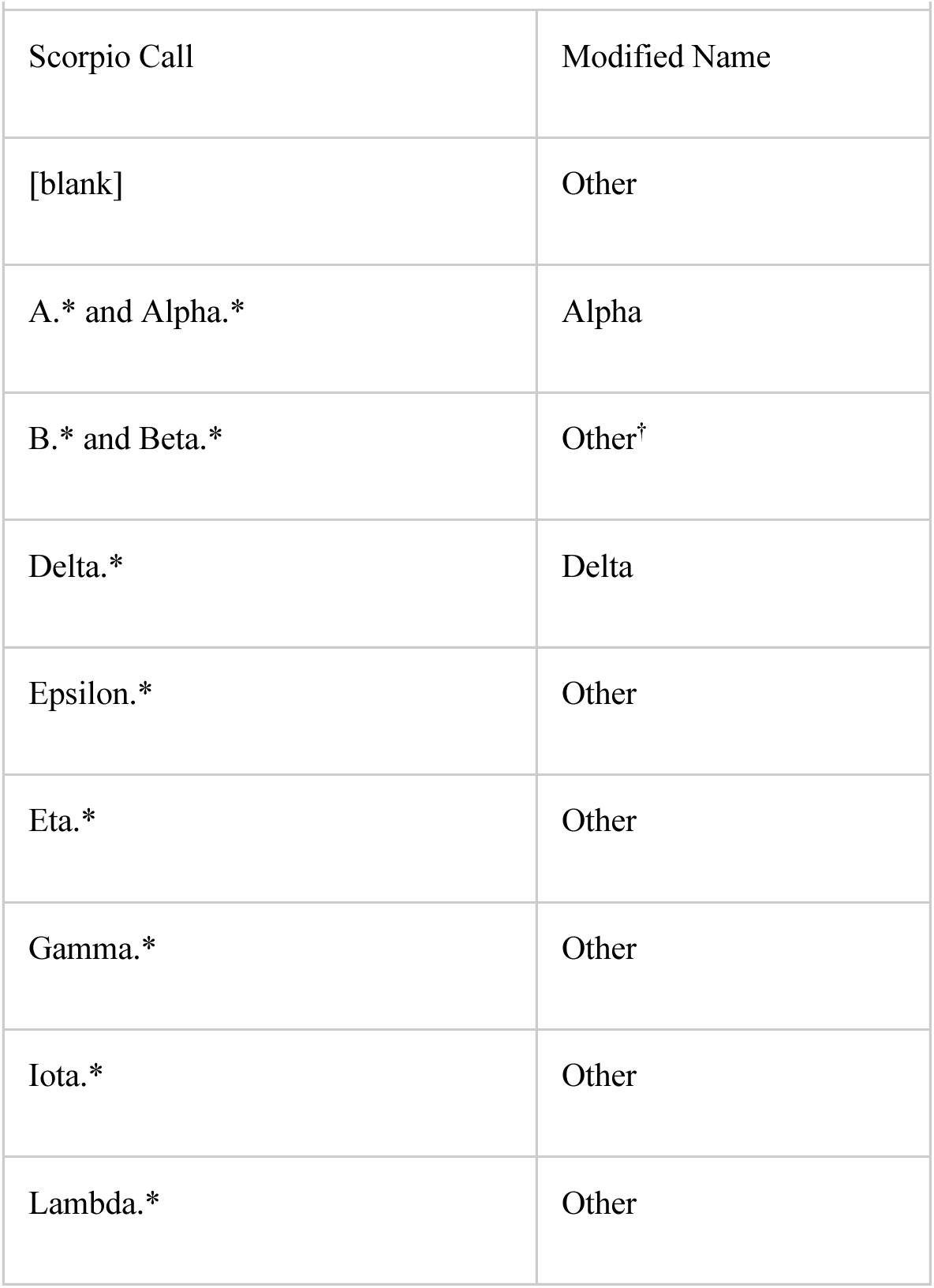
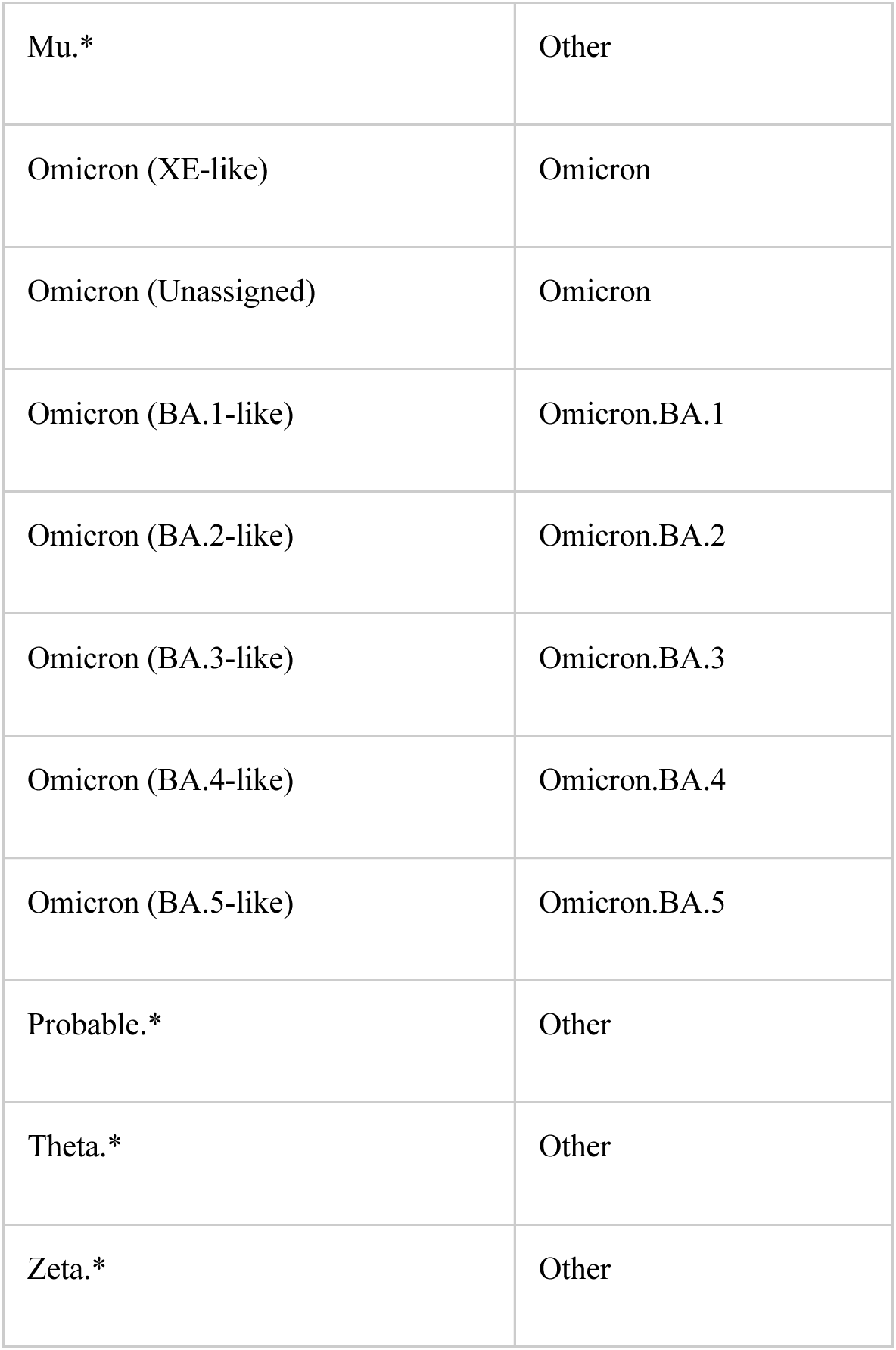
Scorpio Calls.

| Scorpio Call | Modified Name |
| --- | --- |
| [blank] | Other |
| A.* and Alpha.* | Alpha |
| B.* and Beta.* | Other<sup>†</sup> |
| Delta.* | Delta |
| Epsilon.* | Other |
| Eta.* | Other |
| Gamma.* | Other |
| Iota.* | Other |
| Lambda.* | Other |
| Mu.* | Other |
| Omicron (XE-like) | Omicron |
| Omicron (Unassigned) | Omicron |
| Omicron (BA.1-like) | Omicron.BA.1 |
| Omicron (BA.2-like) | Omicron.BA.2 |
| Omicron (BA.3-like) | Omicron.BA.3 |
| Omicron (BA.4-like) | Omicron.BA.4 |
| Omicron (BA.5-like) | Omicron.BA.5 |
| Probable.* | Other |
| Theta.* | Other |
| Zeta.* | Other |

### Pooled Clinical Samples Simulation

We generated survey sequence for each day during the UK pandemic by randomly selecting 100 genomes. We simulated Illumina reads from each of these 100 genomes using wgsim. These reads can stand in for genomes in an analysis of clinical isolates, or they can be combined, scrambled, and surveyed as representative of any one day. We simulated 1 million reads from each sample but kept only 400,000 after all read libraries were joined. These 400,000 reads formed the basis of pairwise KHILL calculations between each day and all days that came before.

### Sketch Depth Comparison

We tested how the sketch depth used during the KHILL calculation impacts the detection of subvariants by calculating KHILL during the course of the UK SARS-CoV-2 pandemic using sketch depths of 100 and 1,000. We used a generalized additive model to fit a cubic spline (lines) with a kurtosis of 50 to these datasets independently in R using the package *mgcv* (https://cran.r-project.org/web/packages/mgcv/index.html). We find that additional genetic diversity is captured with deeper sketch rates, as evidenced by more variation in KHILL during the Delta and Omicron waves (Supplemental Figure 1).

### Diversity Simulations

We simulated mutations in populations of 29.9Kb SARS-CoV-2 genomes using the EMBOSS tool *msbar* (24) and the SARS-CoV-2 reference (NCBI Reference Sequence: NC_045512.2) to understand how numbers of mutations and mutation classes impact the KHill statistic. First, we simulated populations of 1,000 genomes with 1bp SNP mutations. Mutations were simulated to represent 1% - 50% of the genome in steps of 1% (i.e. 299 – 14,952 SNP mutations). Next, we explored how variation in the bock size of mutations (1, 5, 10, 20, 50, 75, 100, 250 and 500 bp) and mutation class impacted the KHILL statistic in populations of 100 genomes. For each block size, we simulated mutation classes: insertion, deletion, SNP (or replacement for block mutations greater than 1 bp), duplication, transposition (copy/paste), or “any”, which is a random combination of all mutation forementioned classes (Supplemental Figure 6).

### KHILL vs. Nucleotide Diversity (π)

We tested how KHILL compares to an existing metric of population genetic diversity, Nei’s nucleotide diversity (π). First, we randomly subsampled 100 SARS-CoV-2 genomes from each day of the UK pandemic and aligned these using *clustal omega* (www.clustal.org). Next, we calculated genome-wide π from each subsample alignment using custom *perl* scripts. For each day, we compared the KHILL measure to the subsample genome-wide π, and fit a linear model to this relationship. Finally, we benchmarked the time required to align 2, 5, 10, 25, 50, 100, 250, 500, and 1,000 randomly subsampled SARS-CoV-2 genomes (from a single day during the UK pandemic) using *clustal omega.* We did this to estimate the computational burden of aligning genomes on the scale sequenced during the UK pandemic. Alignments are required to calculate traditional population genetic measures such as π. It is important to note that these estimates of computational time do not include the time required to: demultiplex and trim raw sequencing data, align this data to a reference genome, generate the consensus sequences used in whole genome alignment, or the calculation of π itself. We sought only to illustrate that when the number of genomes reaches the thousands, the computational time to calculate traditional population genetic measures becomes unmanageable (Supplemental Figure 7).

### Slopes Analysis

We used the R package *mgcv* (https://cran.r-project.org/web/packages/mgcv/index.html) to fit a cubic spine with a kurtosis of 50 to the UK covid pandemic KHILL data using a generalized additive model. We extracted the first derivative F’(x), or slope, of the cubic spline to better understand the rate of change of the spline fit to the KHILL data, and the general trend of the underlying KHILL data during variant sweeps. Positive F’(x) values indicated a positive slope (increasing KHill), while negative F’(x) values indicated a negative slope (decreasing KHill). Rapid shifts in F’(x) mark rapid variant sweeps, especially when the population is dominated by a single variant, and then swept to extinction rapidly by a genetically distant variant (e.g. Delta to Omicron.BA.1 sweep) (Supplemental Figure 8). Sweeps between genetically similar subvariants (e.g. Omicron.BA.1 to Omicron.BA.2) produce more modest fluctuations in F’(x).

## Supplemental Figures Legends

**Supplemental Figure 1.**
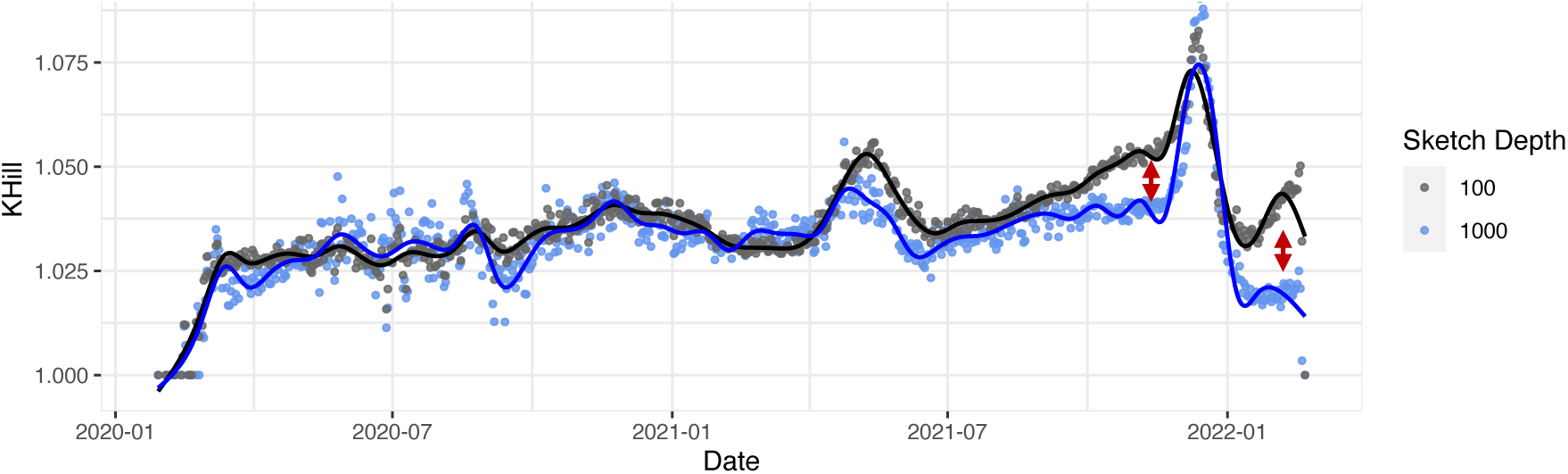
Sketch depth and the detection of subvariants. We used a generalized additive model to fit a cubic spline (lines) with a kurtosis of 50 to the UK covid pandemic KHill statistics (y-axis, as a response to date on the x-axis), calculated with sketch rates of 100 and 1,000. Red arrows indicate the additional diversity captured with a deeper sketch during the Delta and Omicron waves.

**Supplemental Figure 2.**
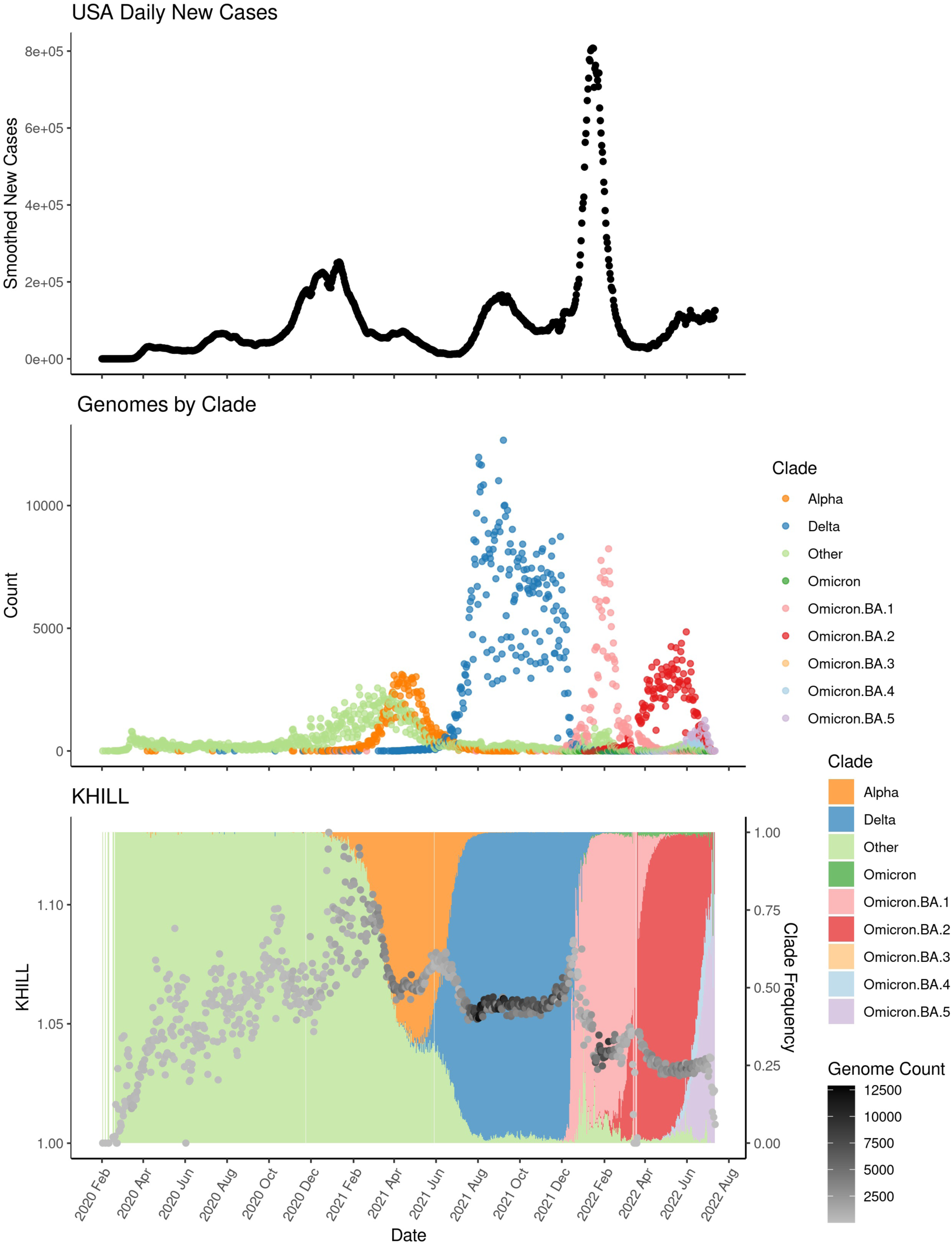
The United States (US) COVID-19 pandemic. We show new case burden (Panel A) and gross phylogenetic classification (Panel B) for the US pandemic since its inception. The arrival of Alpha (March 2021) seems to have homogenized a previously variable US viral population. The Delta surge (June 2021) moderately increases complexity, while the arrival of Omicron (December 2021) is accompanied by a KHILL spike followed by a rapid descent after Delta is forced into near extinction (Panel C).

**Supplemental Figure 3.**
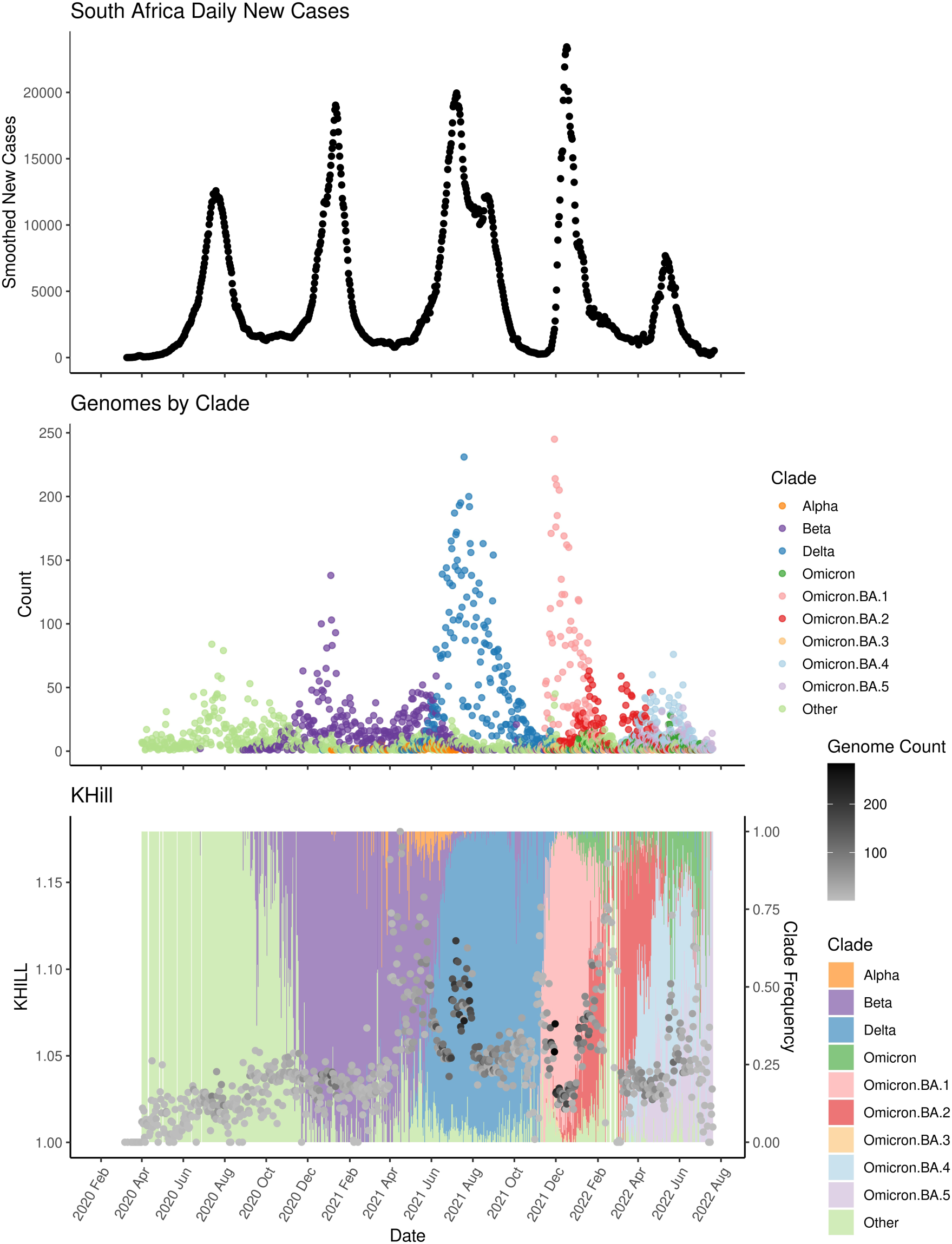
The South Africa (SA) COVID-19 pandemic. We show new case burden (Panel A) and gross phylogenetic classification (Panel B) for the SA pandemic since its inception. The arrival of both Delta and Omicron lead to KHILL spikes. Because of sparser sequencing in this population, the SA data is considerably noisier than either the UK or USA. Note that we also include the Beta variant here as it was a significant variant in the SA pandemic.

**Supplemental Figure 4.**
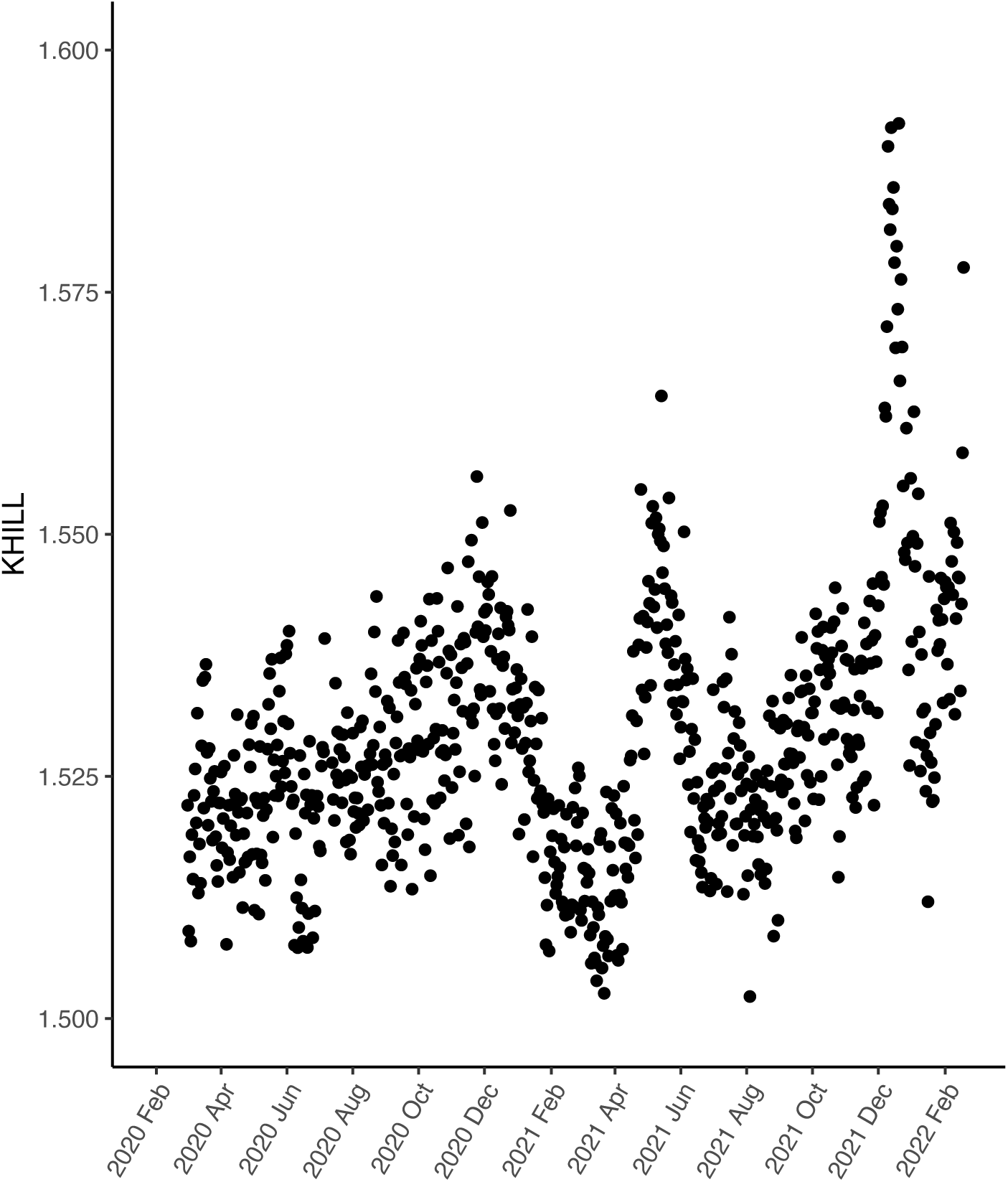
Simulated reads. We simulated 3 Mbases of Illumina sequence from each of 50 randomly selected genomes taken daily over the course of the UK pandemic. We show that the KHILL pandemic curve in this unassembled sequence mirrors that shown in Figure 2 for assembled genomes.

**Supplemental Figure 5.**
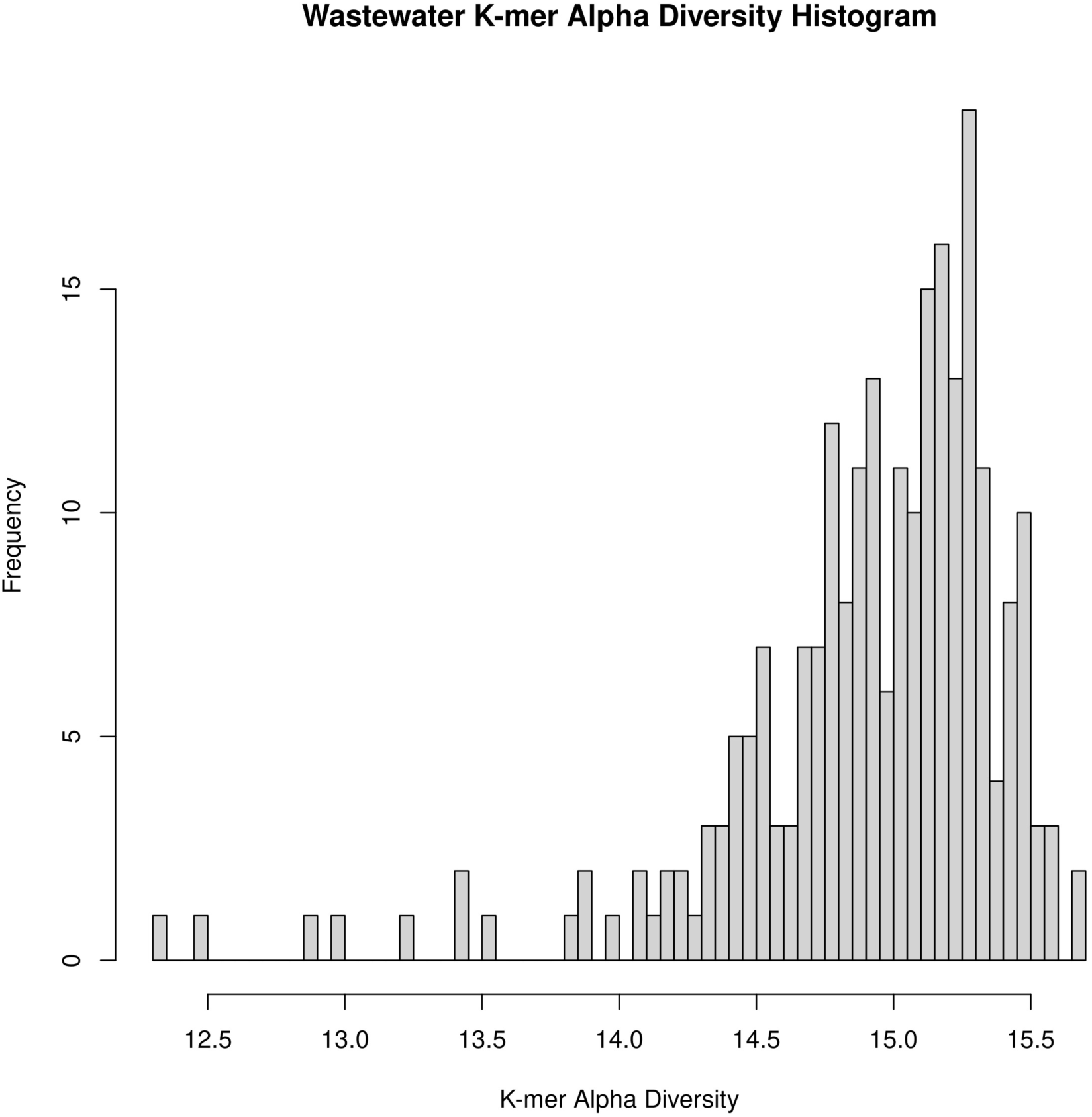
Alpha diversity of wastewater k-mers. We plot the alpha diversity of sequence gathered from each day of the San Diego wastewater sample. Because the sample is amplicon based and enriched for SARS-CoV-2, higher alpha diversity indicates more unique viral k-mers sequenced at a higher depth, while lower diversity is likely characteristic of over-amplified, less even coverage. We remove these low complexity (and some high complexity) days by taking only those samples with sequence one standard deviation from the alpha diversity mean.

**Supplemental Figure 6.**
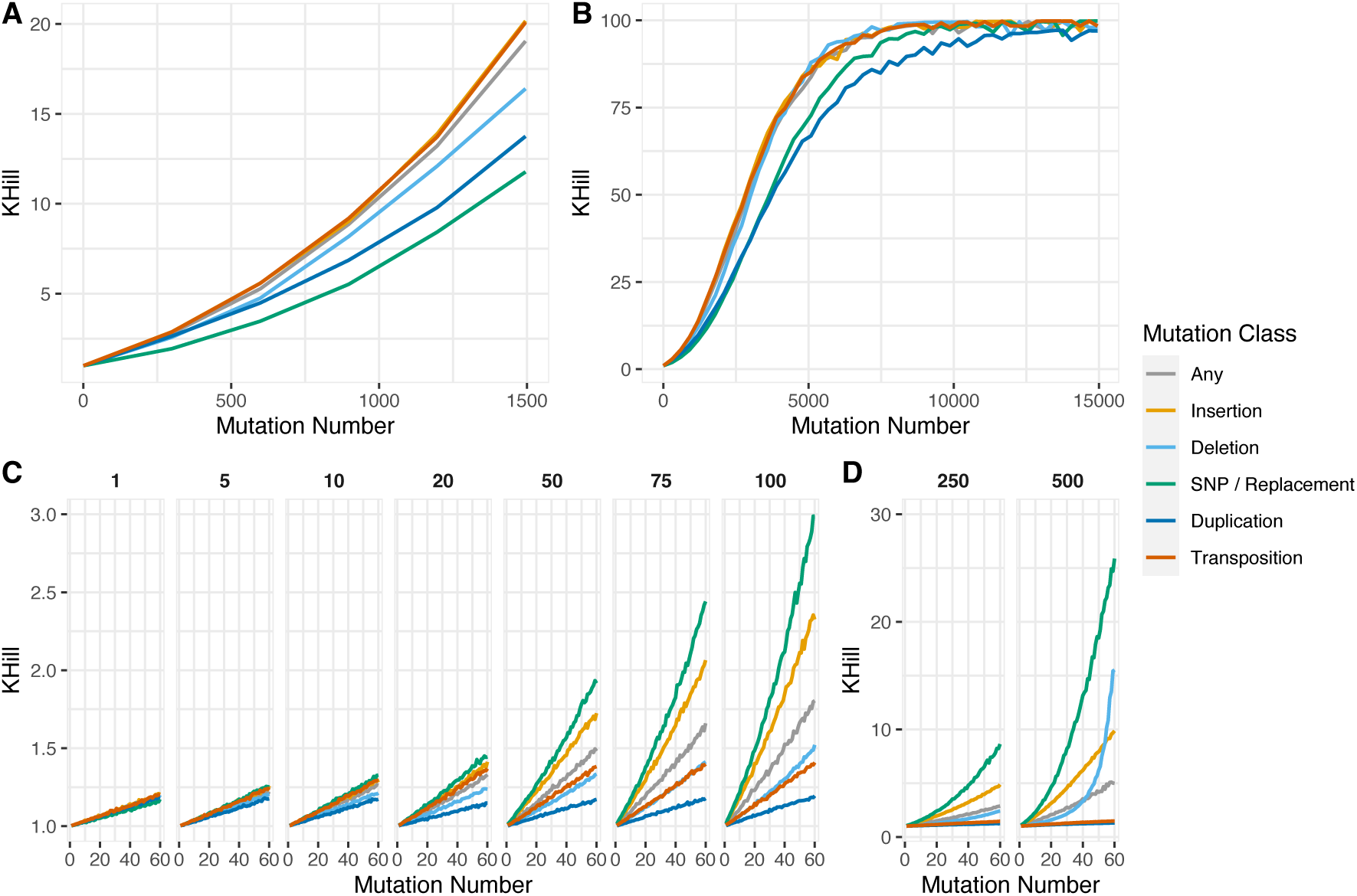
Simulations. We simulated mutations in populations of 29.9Kb SARS-CoV-2 genomes using the EMBOSS tool *msbar* (24) and the SARS-CoV-2 reference (NCBI Reference Sequence: NC_045512.2) to understand how numbers of mutations and mutation classes impact the KHILL statistic. Each population differs in the number of mutations (x-axis), mutation class (legend), and the block size of each mutation (as below). Panels A and B report the KHILL statistic (y-axis) calculated from populations of 1,000 genomes with 1bp SNP mutations. Mutations were simulated to represent 1% - 50% of the genome in steps of 1% (i.e. 299 – 14,952 mutations). Panel A reports the KHILL statistic for zero to 1,500 mutations, while the maximum number of mutations in panel B is 14,952, or 50% of the genome. Panels C and D report KHILL statistics for populations of 100 genomes and are faceted and annotated (top) by mutation block size (1, 5, 10, 20, 50, 75, and 100 bp in panel C, and 250 and 500 bp in panel D). We simulated mutation classes: insertion, deletion, SNP (or replacement for block mutations greater than 1 bp), duplication, transposition (copy/paste), or “any”, which is a random combination of all mutation forementioned classes.

**Supplemental Figure 7.**
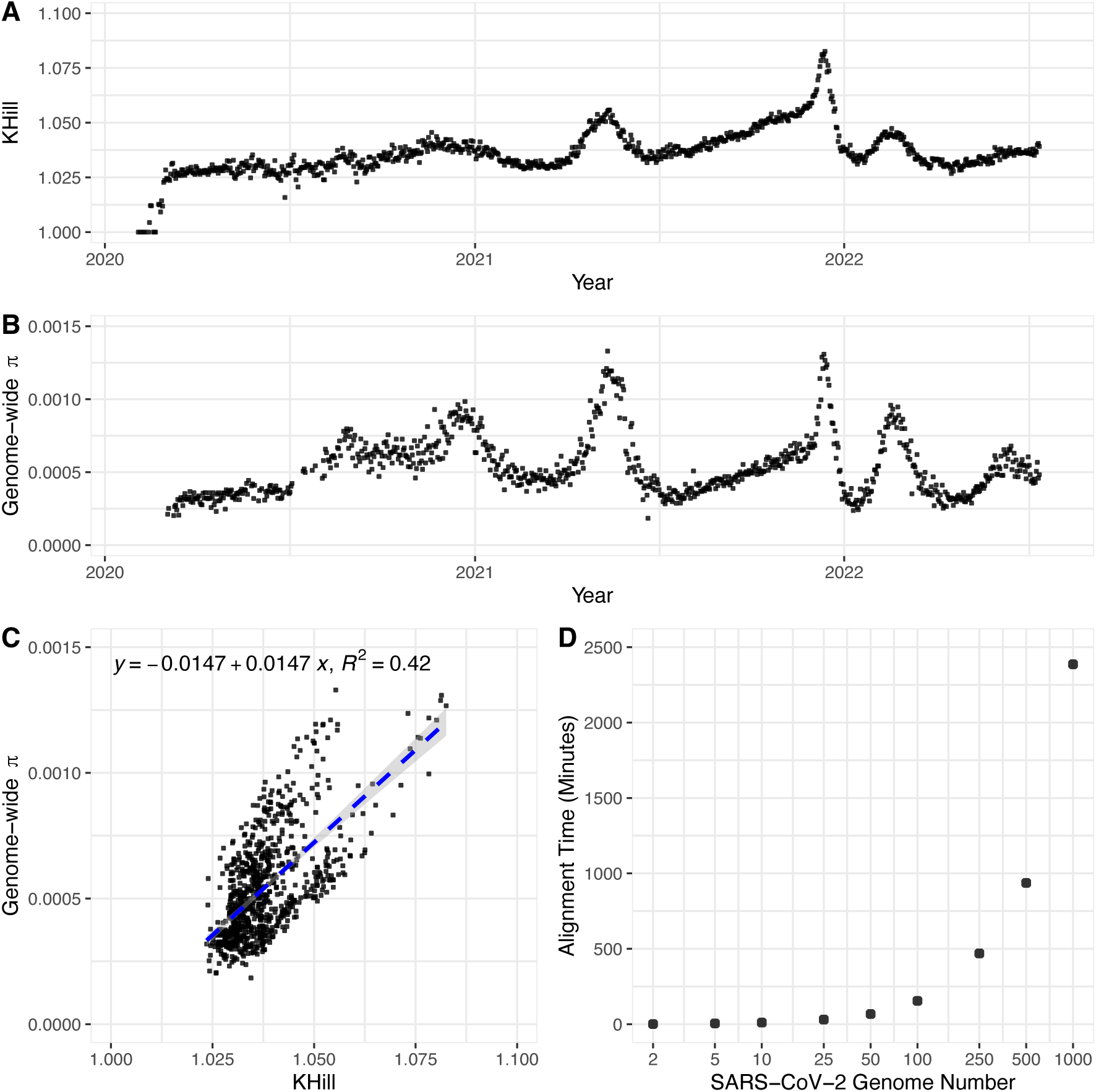
Comparison between KHILL and genome wide nucleotide diversity (π). We show that genome wide π of 100 subsampled genomes per day across the UK pandemic, closely approximates the more data heavy KHILL curve (Panels A and B). Genome wide π is well correlated with KHILL (Panel C), but its calculation is expensive for anything beyond 100 genomes per day because of the cost incurred by whole genome alignment (Panel D).

**Supplemental Figure 8.**
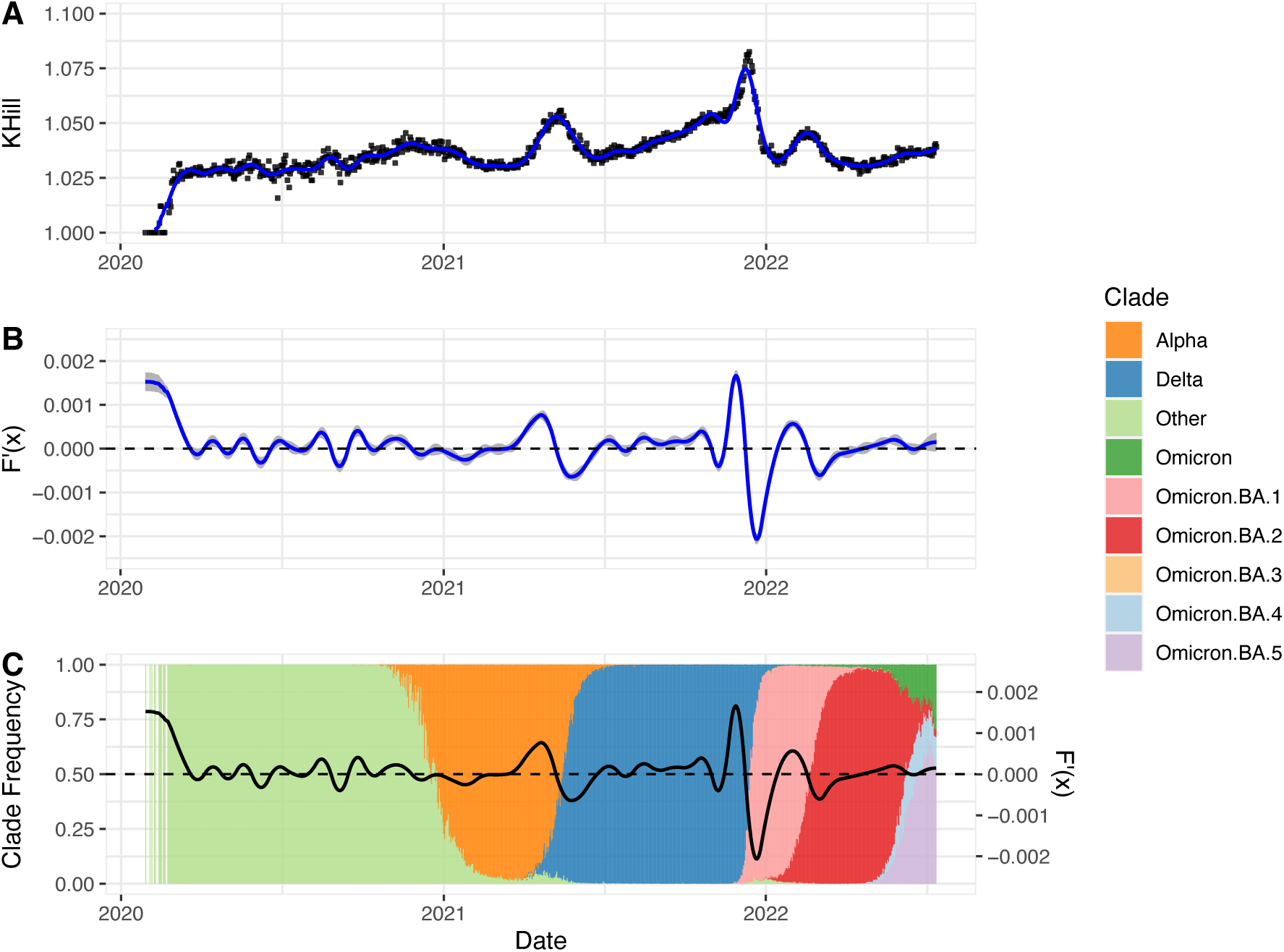
Slopes. In Panel A we used a generalized additive model to fit a cubic spline (blue line) with a kurtosis of 50 to the UK covid pandemic KHill statistics (y-axis, as a response to date on x-axis). Panel B presents the first derivative F’(x), or slope, of the cubic spline presented in Panel A. Positive values indicated a positive slope (increasing KHill), while negative values indicated a negative slope (decreasing KHill). The grey shaded area represents the 95% confidence interval of F’(x). Panel C presents F’(x) (right y-axis) overlayed on the SARS-CoV-2 clade frequency (left y-axis) of the UK covid pandemic dataset. Alpha, Delta, and Omicron sub-variants have been collapsed (legend). In Panel B and C the horizontal dashed line at F’(x) = 0 indicates the transition point from positive to negative slopes of the fitted cubic spline.

